# *Streptococcus pneumoniae* infection is associated with Matrix Metalloproteinase-9 in Lung Cancer Progression and Brain Metastases

**DOI:** 10.1101/2025.04.15.25325903

**Authors:** Lu Gao, Feng Jiang

## Abstract

Matrix metalloproteinases (MMPs) are critical mediators of extracellular matrix remodeling, playing a pivotal role in the progression and metastasis of lung cancer. Emerging evidence indicates that bacterial pathogens, such as Streptococcus pneumoniae (SP), can modulate MMP activity and contribute to tumor progression. This study quantified SP abundance in lung cancer tissues and investigated its relationship with MMP-9 and MMP-2 expression, as well as its impact on clinical outcomes. SP DNA levels were assessed using droplet digital PCR, and MMP-9 and MMP-2 protein expression were evaluated by immunohistochemistry in 120 lung cancer samples. Elevated SP abundance was significantly associated with increased MMP-9 expression, advanced lung cancer stages, greater brain metastases burden, and reduced overall survival (P < 0.05). However, SP abundance showed no correlation with MMP-2 expression. These findings highlight a direct link between SP infection and lung cancer progression through MMP-9–mediated extracellular matrix degradation and metastatic spread. Targeting the SP–MMP-9 axis may represent a novel therapeutic approach to mitigate metastasis and improve patient outcomes in lung cancer.

## Introduction

Lung cancer remains the leading cause of cancer-related mortality worldwide, resulting in millions of deaths each year among both men and women ^1,2^. Approximately 85% of lung cancer cases are classified as non-small cell lung cancer (NSCLC), which primarily includes adenocarcinoma (AC) (40–50%) and squamous cell carcinoma (SCC) (25–30%) ^1,3^. One of the most devastating complications for lung cancer patients is the development of brain metastases (BM), which significantly contribute to mortality and severely diminish quality of life ^4,5^. Despite substantial advances in therapeutic strategies, including targeted therapies and immunotherapies, the molecular mechanisms underlying lung cancer progression and metastasis, particularly to the brain, are not fully understood. Elucidating these mechanisms is crucial for identifying novel therapeutic targets and improving patient outcomes.

Emerging evidence suggests that microbial infections play a significant role in cancer biology ^6^. These infections can contribute to tumor initiation, progression, and metastasis through various mechanisms, including the activation of pro-inflammatory and oncogenic pathways ^6-11^. Previous research from our group has demonstrated that *Streptococcus pneumoniae* (SP), a common respiratory pathogen, binds to lung cancer cells via its pneumococcal surface protein C interacting with the platelet-activating factor receptor ^12,13^. This interaction activates key signaling pathways such as PI3K/AKT and NF-κB, which are known to drive inflammatory responses, tumor proliferation, and progression ^12^. These findings suggest that microbial infections, particularly SP, may actively contribute to the progression of lung cancer and its metastasis to distant organs, including the brain.

Matrix metalloproteinases (MMPs), especially MMP-9 and MMP-2, are critical enzymes involved in extracellular matrix (ECM) remodeling ^14^. ECM serves as a structural barrier, and its degradation is a pivotal step in cancer cell invasion and metastasis. MMP-9 and MMP-2 facilitate the degradation of ECM components such as collagen and gelatin, promoting tumor cell migration and colonization in distant organs, including the brain ^15^. Elevated MMP activity has been strongly associated with tumor progression, metastasis, and poor survival outcomes in cancer patients. Notably, the role of MMP-9 and MMP-2 in the development of brain metastases is of particular interest, given that BM is a leading cause of death in lung cancer patients ^15^. However, the upstream regulators of MMP activity, including potential contributions from microbial pathogens, remain poorly understood ^15^. Given the important role of SP in promoting inflammation and tumor progression, along with the critical involvement of MMP-9 and MMP-2 in cancer metastasis, we hypothesize that SP may promote lung cancer progression and metastasis through pathways involving MMP activity. Specifically, we propose that SP influences MMP-9 and MMP-2 activity, thereby facilitating ECM remodeling, tumor invasion, and metastasis. In this study, we aim to investigate the abundance of SP in lung cancer tissues and its association with MMP-9 and MMP-2 expression, as well as its impact on survival outcomes in lung cancer patients.

## Results

### SP DNA abundance and MMP-9 expression are both markedly elevated in advanced lung cancer

Droplet Digital PCR (ddPCR) analysis shows significantly higher SP DNA levels in advanced-stage tumors compared to early-stage tumors, in patients with brain metastases compared to those without, and in smokers compared to non-smokers (all p<0.05) (Table 1). Patients with SCC have a higher SP abundance compared to those with AC (all p = 0.034). Additionally, higher SP DNA levels is related with positive lymph node involvement, and in those with brain metastases pf lung cancer (all p ≤ 0.05). Gender and age do not significantly influence SP levels (all p > 0.05). Immunohistochemistry was successfully performed on all specimens (Figure 1). IHC analysis revealed that MMP-9 expression was significantly higher in advanced-stage tumors, in SCC compared to adenocarcinoma, in patients with positive lymph node involvement, and in those with brain metastases (all p ≤ 0.05) (Table 1, Figure 2). In contrast, although MMP-2 expression is elevated across all lung cancer tissues, it does not differ significantly between early- and advanced-stage tumors (Figure 2), among various histological types, between smokers and non-smokers, or between patients with and without brain metastases (all p > 0.05) (Table 1).

**Table 1.** SP, MMP-2, and MMP-9 expression and clinical correlations in lung cancer *.

| Variables | Number of Cases (%) | SP (mean ± s.d.) | P-value for SP | MMP-2 (mean ± s.d.) | P-value for MMP-2 | MMP-9 (mean ± s.d.) | P-value for MMP-9 |
| --- | --- | --- | --- | --- | --- | --- | --- |
| **Age (years)** |  |  |  |  |  |  |  |
| >69 | 49 (40.8%) | 3.56 ± 1.102 | | 38.2 ± 10.5 | | 56.8 ± 12.3 | |
| <69 | 71 (59.2%) | 2.83 ± 0.85 | 0.205 | 36.7 ± 11.2 | 0.331 | 53.6 ± 11.5 | 0.267 |
| **Gender** |  |  |  |  |  |  |  |
| Male | 87 (72.5%) | 3.37 ± 0.98 | | 39.4 ± 10.3 | | 57.4 ± 11.7 | |
| Female | 33 (27.5%) | 2.94 ± 0.89 | 0.165 | 35.8 ± 11.4 | 0.325 | 54.1 ± 12.0 | 0.059 |
| **Histology** |  |  |  |  |  |  |  |
| AC | 62 (51.7%) | 2.98 ± 0.76 | | 36.1 ± 9.8 | | 52.3 ± 11.2 | |
| SCC | 58 (48.3%) | 3.62 ± 1.104 | 0.005 | 40.5 ± 10.7 | 0.328 | 60.2 ± 12.5 | 0.034 |
| **Smoking Status** |  |  |  |  |  |  |  |
| Smokers | 104 (86.7%) | 3.45 ± 0.93 | | 39.0 ± 10.4 | | 58.6 ± 12.0 | |
| Non-smokers | 16 (13.3%) | 2.78 ± 0.79 | 0.041 | 35.6 ± 9.9 | 0.312 | 54.8 ± 11.3 | 0.058 |
| **T-status** |  |  |  |  |  |  |  |
| 1a | 24 (20.0%) | 3.06 ± 0.82 | | 35.9 ± 10.1 | | 54.2 ± 11.1 | |
| 1b | 16 (13.3%) | 3.11 ± 0.87 | | 36.3 ± 9.8 | | 55.3 ± 11.8 | |
| 2 | 50 (41.7%) | 3.32 ± 0.91 | | 38.4 ± 10.5 | | 57.4 ± 11.6 | |
| 3 | 21 (17.5%) | 3.64 ± 1.00 | | 40.2 ± 11.2 | | 59.6 ± 12.4 | |
| 4 | 9 (7.5%) | 3.79 ± 1.12 | 0.017 | 41.8 ± 10.9 | 0.321 | 61.2 ± 12.7 | 0.028 |
| **N-status** |  |  |  |  |  |  |  |
| No | 48 (40.0%) | 2.87 ± 0.79 | | 36.0 ± 9.6 | | 52.6 ± 11.3 | |
| Yes | 72 (60.0%) | 3.58 ± 1.05 | 0.013 | 39.8 ± 10.4 | 0.316 | 58.9 ± 12.2 | 0.016 |
| **Brain Metastases** |  |  |  |  |  |  |  |
| Absent | 100 (83.3%) | 3.02 ± 0.89 | | 36.5 ± 10.0 | | 54.7 ± 11.6 | |
| Present | 20 (16.7%) | 3.78 ± 1.12 | 0.011 | 39.4 ± 10.8 | 0.319 | 61.3 ± 12.5 | 0.026 |
| **Stage** |  |  |  |  |  |  |  |
| I | 32 (26.7%) | 2.79 ± 0.68 | | 35.2 ± 9.7 | 0.308 | 51.9 ± 10.9 | |
| II | 57 (47.5%) | 3.53 ± 0.94 | | 38.7 ± 10.2 | 0.315 | 57.2 ± 12.0 | |
| III | 31 (25.8%) | 3.87 ± 1.06 | 0.028 | 40.9 ± 10.8 | 0.314 | 60.7 ± 12.6 | 0.012 |

\*, Statistical differences were evaluated using the Mann-Whitney U test for comparisons between two groups and the Kruskal-Wallis test for comparisons among multiple groups.

**Figure 1.**
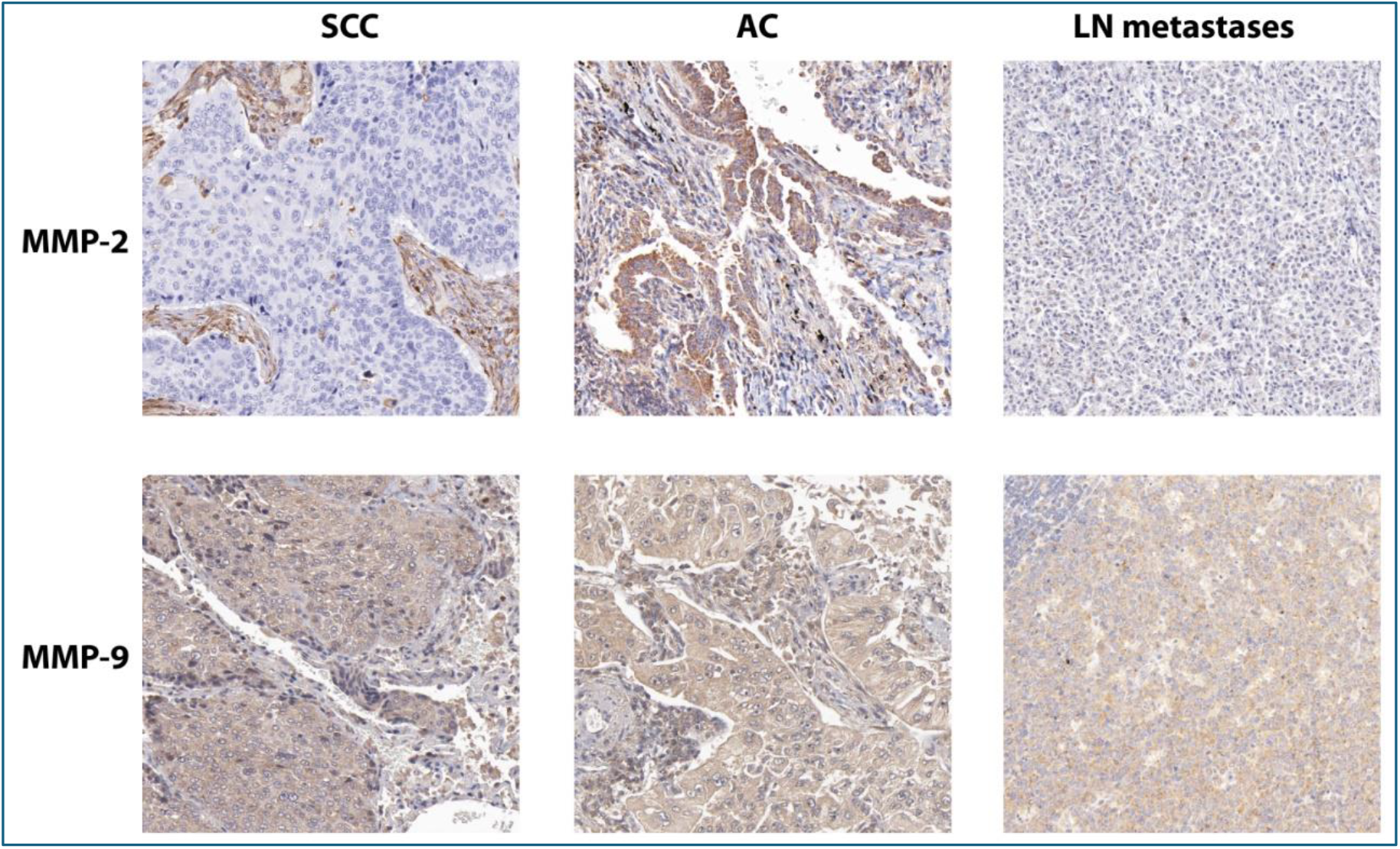
IHC staining of MMP-2 and MMP-9 in lung cancer subtypes and lymph node (LN) metastases. The top row shows MMP-2 staining with moderate positivity in squamous cell carcinoma (SCC) and adenocarcinoma (AC), while minimal expression is observed in LN metastases. The bottom row highlights strong MMP-9 staining across SCC, AC, and LN metastases, indicating elevated expression in these tissues. Magnification: 200x.

**Figure 2.**
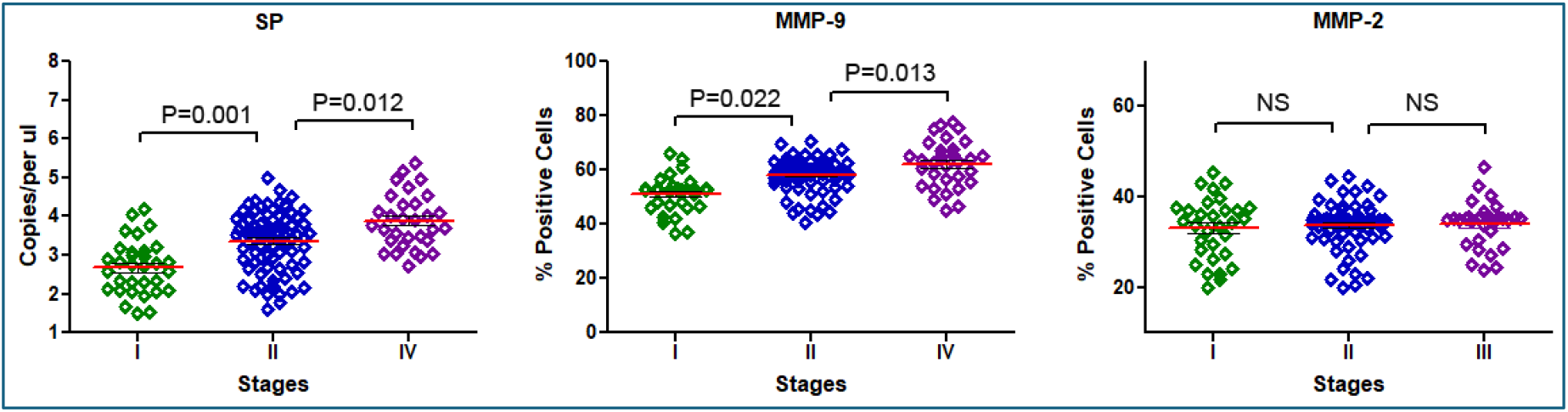
The expression levels of SP, MMP-2, and MMP-9 across lung cancer stages I, II, and III. SP expression, represented by the leftmost bar in each group, shows a significant increase with advancing cancer stage, accompanied by error bars indicating standard deviations. MMP-9 expressions similarly rise markedly from stage I to stage III, highlighting its strong association with cancer progression and metastasis. In contrast, MMP-2 expression remains relatively stable across all stages. The y-axis reflects expression levels or the percentage of positive cells, and the error bars denote standard deviations.

### SP Abundance Correlates with MMP-9 Expression, Tumor Characteristics, and Tumor Grades

A positive correlation was observed between SP abundance and MMP-9 expression (all p < 0.05) across tumor stages (Table 2) (Figure 3). In contrast, SP abundance did not correlate significantly with MMP-2 expression across tumor stages (p > 0.05) (Table 2, Figure 3). SP abundance demonstrated a significant correlation with T-status, particularly in higher T-status categories. In T3 (Spearman’s ρ = 0.72, p = 0.008) and T4 tumors (Spearman’s ρ = 0.78, p = 0.004). Positive correlations were also observed between SP abundance and N-status (Spearman’s ρ = 0.67, p = 0.003), reflecting its association with lymph node involvement. A stage-dependent pattern of SP abundance was evident, with significant correlations in Stage II (Spearman’s ρ = 0.62, p = 0.017) and Stage III (Spearman’s ρ = 0.71, p = 0.009) (Table 2, Figure 3). Additionally, SP abundance exhibited significant correlations with tumor grade, showing stronger correlations in intermediate-grade (Spearman’s ρ = 0.51, p = 0.041) and high-grade tumors (Spearman’s ρ = 0.68, p = 0.002), highlighting its relationship with aggressive tumor phenotypes. SP abundance was also significantly correlated with brain metastases, further emphasizing the role of SP in advanced disease and metastasis. These results confirm the role of SP in lung cancer progression, aggressive disease phenotypes, and metastatic potential, providing a comprehensive perspective on its relationship with tumor characteristics and grades.

**Table 2:**
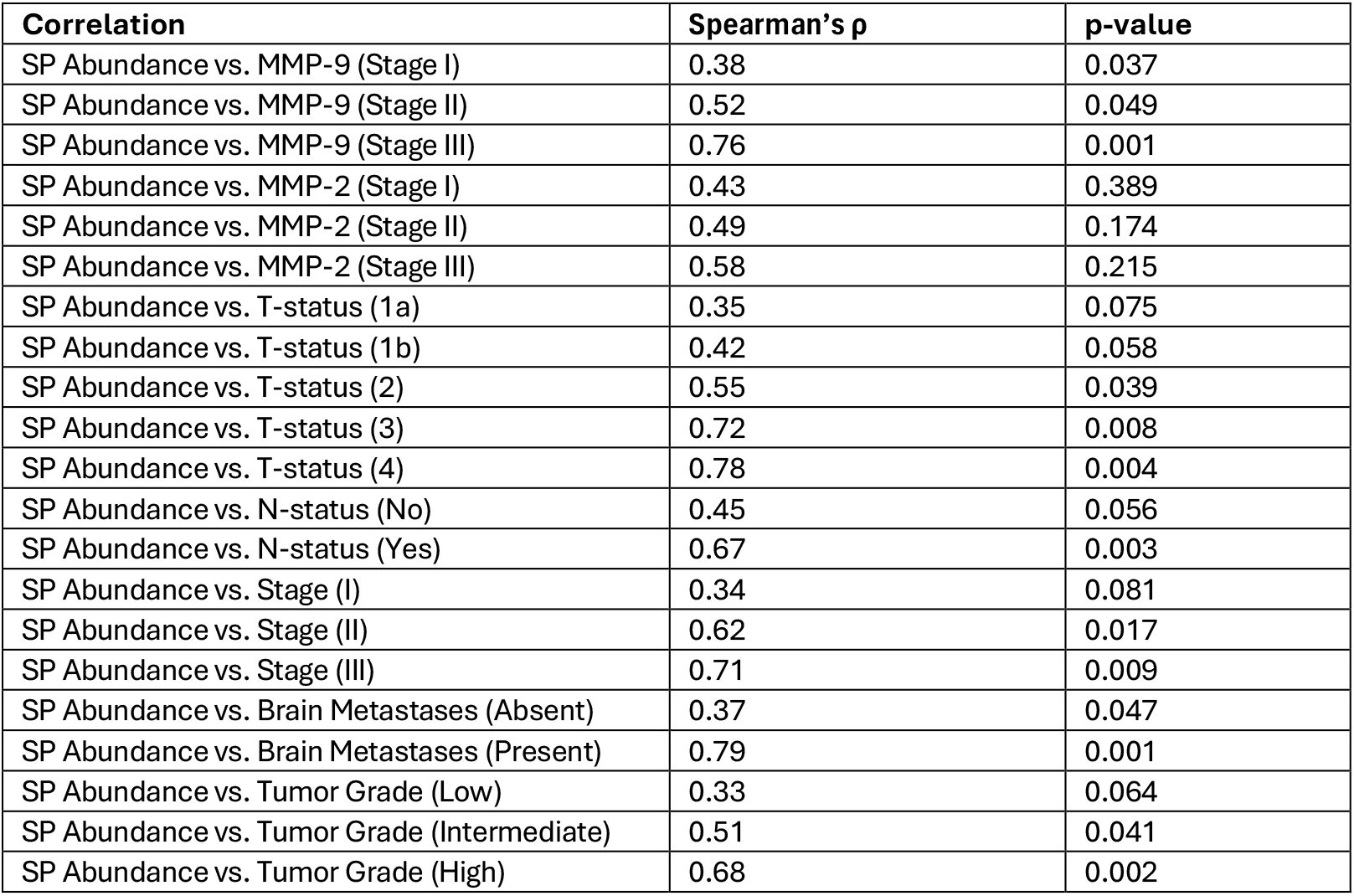
Association of SP levels with MMPs and lung tumor features.

| Correlation | Spearman's $\rho$ | p-value |
| --- | --- | --- |
| SP Abundance vs. MMP-9 (Stage I) | 0.38 | 0.037 |
| SP Abundance vs. MMP-9 (Stage II) | 0.52 | 0.049 |
| SP Abundance vs. MMP-9 (Stage III) | 0.76 | 0.001 |
| SP Abundance vs. MMP-2 (Stage I) | 0.43 | 0.389 |
| SP Abundance vs. MMP-2 (Stage II) | 0.49 | 0.174 |
| SP Abundance vs. MMP-2 (Stage III) | 0.58 | 0.215 |
| SP Abundance vs. T-status (1a) | 0.35 | 0.075 |
| SP Abundance vs. T-status (1b) | 0.42 | 0.058 |
| SP Abundance vs. T-status (2) | 0.55 | 0.039 |
| SP Abundance vs. T-status (3) | 0.72 | 0.008 |
| SP Abundance vs. T-status (4) | 0.78 | 0.004 |
| SP Abundance vs. N-status (No) | 0.45 | 0.056 |
| SP Abundance vs. N-status (Yes) | 0.67 | 0.003 |
| SP Abundance vs. Stage (I) | 0.34 | 0.081 |
| SP Abundance vs. Stage (II) | 0.62 | 0.017 |
| SP Abundance vs. Stage (III) | 0.71 | 0.009 |
| SP Abundance vs. Brain Metastases (Absent) | 0.37 | 0.047 |
| SP Abundance vs. Brain Metastases (Present) | 0.79 | 0.001 |
| SP Abundance vs. Tumor Grade (Low) | 0.33 | 0.064 |
| SP Abundance vs. Tumor Grade (Intermediate) | 0.51 | 0.041 |
| SP Abundance vs. Tumor Grade (High) | 0.68 | 0.002 |

**Figure 3.**
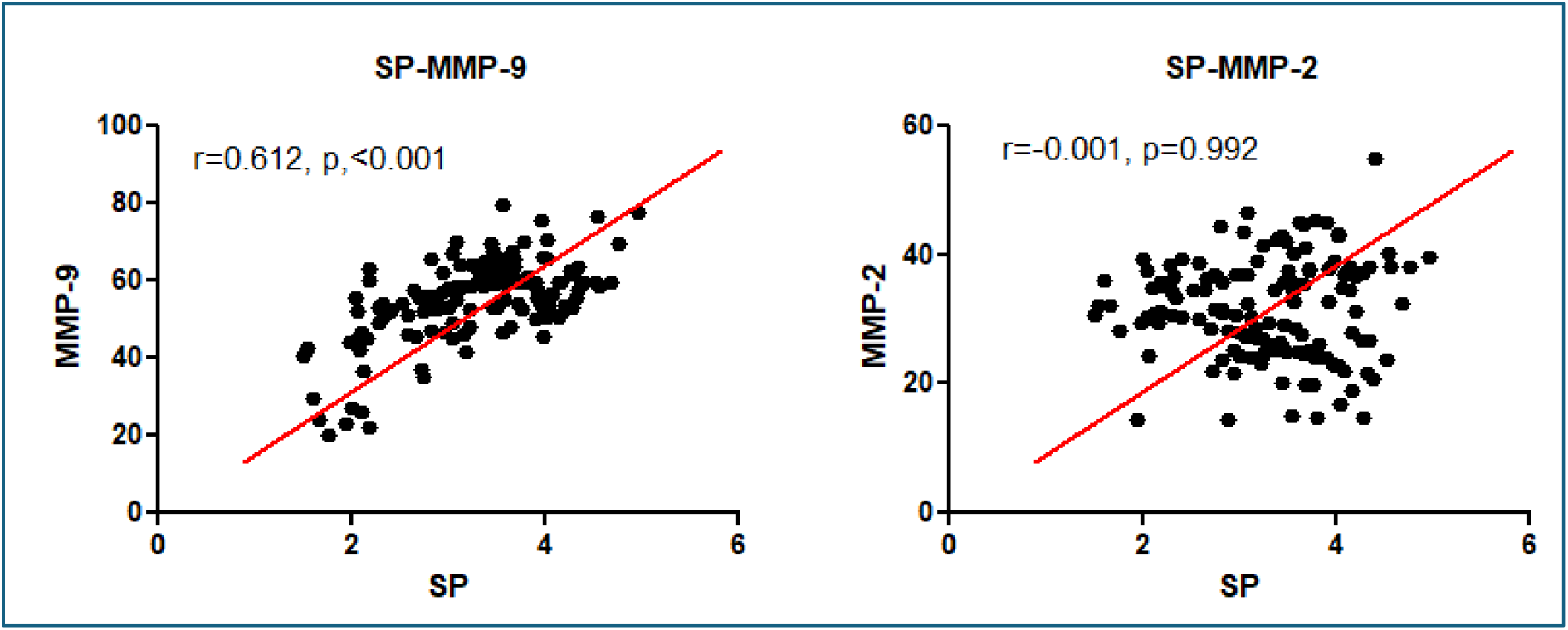
Scatter plots show the correlation (Pearson’s correlation analysis) between SP levels (x-axis) and MMP-9 (left panel) or MMP-2 (right panel) levels (y-axis). In the left panel, a significant positive correlation was observed between SP and MMP-9 (r = 0.612, p < 0.001), as indicated by the upward trend of the data and the fitted regression line (red). In contrast, the right panel shows no significant correlation between SP and MMP-2 (r = -0.001, p = 0.992), with data points scattered randomly around the regression line.

### SP abundance and MMP-9 expression as predictors of survival outcomes in cancer patients

A Kaplan-Meier survival analysis revealed that patients with high SP abundance had significantly shorter overall survival (OS) compared to those with low SP levels (p < 0.01) (Figure 4). Multivariate Cox regression analysis confirmed that SP abundance and MMP-9 expression were independent predictors of poorer survival outcomes (hazard ratio [HR] for SP: 2.1, p = 0.02; HR for MMP-9: 2.8, p < 0.01) (Table 3).

**Figure 4.**
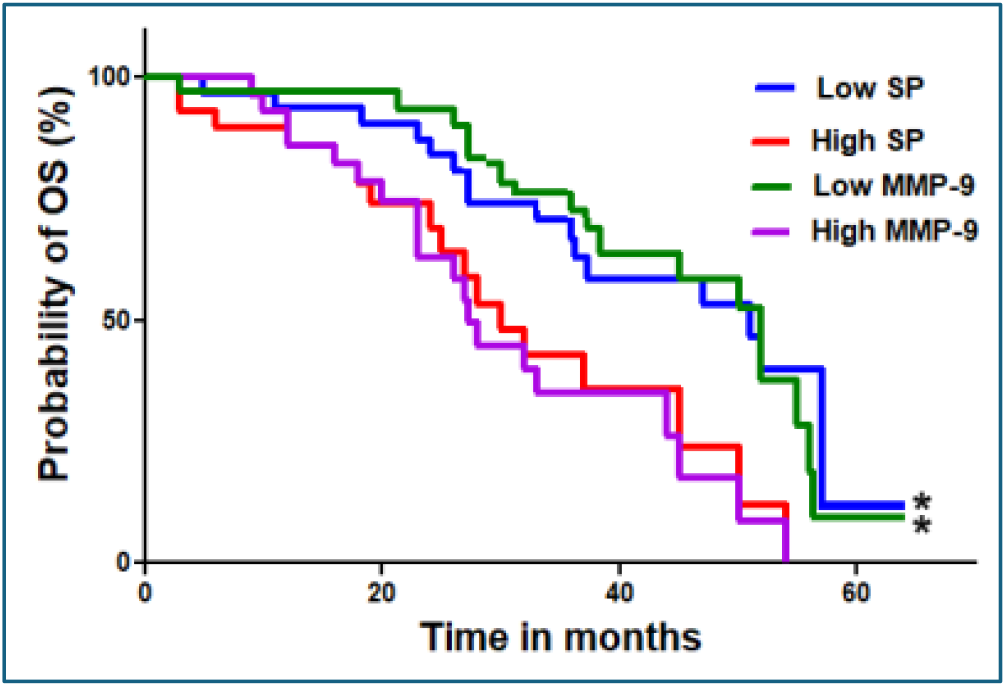
Kaplan–Meier survival curves showing the probability of overall survival (OS) over time (in months) for patients stratified by SP and MMP-9 levels. Patients with low SP (blue line) and low MMP-9 (green line) show improved survival probabilities compared to those with high SP (red line) and high MMP-9 (magenta line). Median OS was 12 months (p < 0.01) in this analysis. Significant differences in survival outcomes between groups are indicated by an asterisk (*), suggesting that high SP and high MMP-9 levels are associated with worse overall survival.

**Table 3.** Associations of SP abundance, MMP-9, and MMP-2 levels with clinical outcomes in lung cancer.

| Clinical Outcome | High SP Abundance | High MMP-9 Levels | High MMP-2 Levels | Low SP Abundance |
| --- | --- | --- | --- | --- |
| Median Overall Survival (OS) | 12 months (p < 0.01) |  | No association (p > 0.05) | 20 months |
| Hazard Ratio (HR) for SP | 2.1 (p = 0.02) |  |  |  |
| Hazard Ratio (HR) for MMP-9 |  | 2.8 (p < 0.01) |  |  |
| Hazard Ratio (HR) for MMP-2 |  |  |  |  |

Kaplan-Meier survival analysis was used to estimate median overall survival (OS), with differences between groups compared using the log-rank test. Hazard ratios (HR) for SP abundance, MMP-9, and MMP-2 levels were calculated using Cox proportional hazards regression, with corresponding p-values indicating the significance of associations between these variables and clinical outcomes.

## Discussion

In this study, we elucidated the role of the bacterial pathogen SP in lung cancer progression and metastasis, emphasizing its association with MMPs. Our findings demonstrate a strong positive correlation between SP abundance and MMP-9 levels. Furthermore, both SP abundance and MMP-9 levels are significantly associated with advanced lung cancer stages, brain metastases, and poor survival outcomes. We also confirmed that SP abundance is associated with smoking status, supporting our previous findings and highlighting smoking as a potential enhancer of SP-related pathways in tumor progression ^12,13^. Additionally, SP abundance was more strongly associated with SCC than AC, implying that SP may play a subtype-specific role in lung cancer pathogenesis. These results underscore the clinical significance of the SP–MMP-9 axis and highlight its potential as a therapeutic target, particularly for smokers at high risk of developing aggressive lung cancer phenotypes.

Interestingly, MMP-2 expression did not exhibit a significant association with advanced lung cancer stages, brain metastases, or survival outcomes. This suggests that MMP-2 may play a distinct and possibly less critical role in SP-mediated tumor biology compared to MMP-9. These findings emphasize that not all MMPs contribute equally to cancer progression and underscore the specificity of SP’s interaction with MMP-9 in promoting metastatic disease. Collectively, our study suggests that microbial infections like SP may play a critical role in modulating the tumor microenvironment, facilitating metastasis, and influencing patient survival outcomes.

The pronounced association between SP abundance and brain metastases is particularly noteworthy, given that brain metastases are a leading cause of mortality among lung cancer patients and their underlying mechanisms remain inadequately understood^1^. Our data propose that SP-driven MMP-9 activity may be instrumental in promoting brain metastases through enhanced tumor invasiveness and ECM degradation. This introduces a new perspective on the metastatic processes in lung cancer, implicating microbial influences in tumor progression and raising important questions about the role of infections. From a clinical standpoint, our findings may open promising avenues for therapeutic intervention. Targeting SP colonization in lung cancer patients could potentially disrupt the SP–MMP-9 axis, thereby reducing ECM degradation and metastatic spread.

Moreover, MMP-9 inhibitors, which are currently under investigation for various malignancies^16^, may offer heightened efficacy in patients with elevated SP levels. The prospective combination of antimicrobial therapies with MMP-9 inhibitors presents an innovative strategy to mitigate metastasis and improve survival outcomes. This study also underscores the broader importance of tumor–microbe interactions in cancer biology.

While significant emphasis has been placed on immune checkpoint inhibition and tumor-intrinsic pathways, our results highlight the substantial impact that microbial infections can have on shaping the tumor microenvironment and influencing therapeutic responses. Future research should aim to further elucidate the interplay between microbial pathogens, MMP activity, and immune modulation to identify additional therapeutic targets.

Nevertheless, our study has some limitations. The reliance on tissue specimens from a single institution may limit the generalizability of our findings; therefore, larger, multicenter studies are necessary to validate and extend these results. Furthermore, while we have established significant correlations between SP abundance, MMP-9 expression, and clinical outcomes, the precise mechanistic pathways involved require further elucidation through experimental models.

In conclusion, our findings provide compelling evidence that SP abundance significantly contributes to lung cancer progression and brain metastases via MMP-9–mediated ECM remodeling. These results not only highlight the importance of microbial infections in cancer biology but also offer promising avenues for therapeutic intervention. Future studies focusing on the SP–MMP-9 interaction may lead to novel strategies aimed at improving outcomes for patients with lung cancer.

## Materials and Methods

### Patients and clinical specimens

The study was conducted under protocols approved by the Institutional Review Board of the University of Maryland Baltimore (IRB HP-00040666), and all patients provided written informed consent. We obtained formalin-fixed, paraffin-embedded (FFPE) lung cancer tissues of 126 NSCLC patients who had either a lobectomy or a pneumonectomy at University of Maryland Medical Center. Clinical and pathological data, including patient age, sex, smoking status, tumor stage, histological subtype, and presence of brain metastases, were obtained from medical records.

### Droplet digital PCR (ddPCR)

SP DNA abundance was quantified using ddPCR. Genomic DNA was extracted from FFPE specimens using the QIAamp DNA FFPE Tissue Kit (QIAGEN, Germantown, MD) as previously described ^12,13^. DNA quality and concentration were assessed using a NanoDrop spectrophotometer (Thermo Fisher Scientific, Waltham, MA). ddPCR was performed using the QX200 System (Bio-Rad Laboratories, Hercules, CA) with specific primers and probes targeting SP genes as previously described ^12,13^. Each reaction mixture contained 10 ng of template DNA, primers and probes at optimal concentrations, and 2X Supermix (Bio-Rad). Droplets were generated using the QX200 Droplet Generator, amplified in a thermal cycler under standard cycling conditions, and analyzed using the QX200 Droplet Reader. Results were reported as copies of SP DNA per microliter, normalized to the total DNA input.

Negative controls (no-template DNA) and positive controls (SP genomic DNA) were included in all experiments.

### Immunohistochemistry (IHC) and analysis

IHC analysis of MMP-2 and MMP-9 was performed on FFPE tissue sections as previously described ^17^. Sections underwent antigen retrieval, followed by incubation with primary antibodies specific to MMP-2 and MMP-9 (Abcam, Waltham, MA). Subsequently, sections were treated with secondary antibodies, and chromogen development was carried out using DAB (Abcam), followed by counterstaining with hematoxylin. Visualization was performed using an Olympus microscope (Olympus. Center Valley, PA). For quantitative analysis, five representative regions encompassing both cellular and stromal areas were selected. Positive cells exhibiting distinct, strong brown staining were manually counted using AnalySIS Pro software (Olympus). To ensure objectivity, counts were independently performed three times per region by two staff members. Results were classified on a scale of 0 (0–10% positive cells) to 3 (>50% positive cells). Image analysis was conducted in a blind manner to eliminate bias.

### Statistical analysis

Statistical analyses were performed to assess relationships between SP abundance, MMP expression, clinical features, and survival outcomes. Descriptive statistics were used to summarize patient demographics, tumor characteristics, and molecular data. Differences in SP abundance and MMP expression across clinical subgroups were analyzed using the Mann-Whitney U test or Kruskal-Wallis test for non-parametric data. Correlations between SP DNA levels, MMP-9, and MMP-2 expression were evaluated using Spearman’s rank correlation coefficient. Survival analyses were conducted using Kaplan-Meier curves and compared using the log-rank test. OS was calculated from the date of diagnosis to the date of death or last follow-up. Cox proportional hazards regression models were used to identify independent predictors of survival, adjusting for potential confounders such as age, sex, tumor stage, and histological subtype. All statistical analyses were performed using SPSS (version 30) and R with a significant level set at p < 0.05. Data visualization was conducted using GraphPad Prism (GraphPad Software, La Jolla, CA).

## Data Availability

All data produced in the present study are available upon reasonable request to the authors

## Author contributions

L. G and F. J. performed experiments and wrote the main manuscript. F.J. developed the main project idea and conceptual framework. All authors reviewed the manuscript.

## Declarations

### Informed consent and approval

The study was conducted under protocols approved by the Institutional Review Board of the University of Maryland Baltimore (IRB HP-00040666), and all patients provided written informed consent.

### Additional information and Data Availability

The data that support the findings of this study are available from the corresponding author upon reasonable request.

## Acknowledgements

The authors wish to thank the Biostatistics Shared Service at the University of Maryland Marlene and Stewart Greenebaum Cancer Center for their invaluable contribution in conducting the statistical analysis for this study.

## References

1. Guan, C., Zhang, X. & Yu, L. A Review of Recent Advances in the Molecular Mechanisms Underlying Brain Metastasis in Lung Cancer. Mol Cancer Ther 23, 627–637 (2024).

2. Leiter, A., Veluswamy, R.R. & Wisnivesky, J.P. The global burden of lung cancer: current status and future trends. Nat Rev Clin Oncol 20, 624–639 (2023).

3. Mithoowani, H. & Febbraro, M. Non-Small-Cell Lung Cancer in 2022: A Review for General Practitioners in Oncology. Curr Oncol 29, 1828–1839 (2022).

4. Hendriks, L.E.L., Cadranel, J. & Berghmans, T. Current challenges in the management of nonsmall cell lung cancer brain metastases. Eur Respir J 55(2020).

5. Souza, V.G.P., et al. Advances in the Molecular Landscape of Lung Cancer Brain Metastasis. Cancers (Basel) 15(2023).

6. Garrett, W.S. Cancer and the microbiota. Science 348, 80–86 (2015).

7. Mao, Q., et al. Interplay between the lung microbiome and lung cancer. Cancer Lett 415, 40–48 (2018).

8. Wang, K., et al. A Preliminary Study of Microbiota Diversity in Saliva and Bronchoalveolar Lavage Fluid from Patients with Primary Bronchogenic Carcinoma. Med Sci Monit 25, 2819–2834 (2019).

9. Tsay, J.J., et al. Lower Airway Dysbiosis Affects Lung Cancer Progression. Cancer Discov 11, 293–307 (2021).

10. Tsay, J.J., et al. Airway Microbiota Is Associated with Upregulation of the PI3K Pathway in Lung Cancer. Am J Respir Crit Care Med 198, 1188–1198 (2018).

11. Tsay, J.C., et al. Molecular characterization of the peripheral airway field of cancerization in lung adenocarcinoma. PLoS One 10, e0118132 (2015).

12. Li, N., et al. Streptococcus pneumoniae promotes lung cancer development and progression. iScience 26, 105923 (2023).

13. Leng, Q., Holden, V.K., Deepak, J., Todd, N.W. & Jiang, F. Microbiota Biomarkers for Lung Cancer. Diagnostics (Basel) 11(2021).

14. Winkler, J., Abisoye-Ogunniyan, A., Metcalf, K.J. & Werb, Z. Concepts of extracellular matrix remodelling in tumour progression and metastasis. Nat Commun 11, 5120 (2020).

15. Wang, Y., et al. Tumor Immune Microenvironment and Immunotherapy in Brain Metastasis From Non-Small Cell Lung Cancer. Front Immunol 13, 829451 (2022).

16. Rashid, Z.A. & Bardaweel, S.K. Novel Matrix Metalloproteinase-9 (MMP-9) Inhibitors in Cancer Treatment. Int J Mol Sci 24(2023).

17. Jiang, F., et al. Aldehyde dehydrogenase 1 is a tumor stem cell-associated marker in lung cancer. Mol Cancer Res 7, 330–338 (2009).

